# ARIMA modelling of predicting COVID-19 infections

**DOI:** 10.1101/2020.04.18.20070631

**Authors:** W.Regis Anne, S.Carolin Jeeva

## Abstract

The World Health Organization (WHO) Director-General, Dr. Tedros Adhanom Ghebreyesus on March 11, 2020 declared the novel coronavirus (COVID-19) outbreak a global pandemic [4] the reason being the number of cases outside China increased 13-fold and the number of countries with cases increased threefold. In this paper a time series model to predict short-term prediction of the transmission of the exponentially growing COVID-19 time series is modelled and studied. Auto Regressive Integrated Moving Average (ARIMA) model prediction is performed on the number of cumulative cases over a time period and is validated over Akaike information criterion (AIC) statistics.

## 1. Introduction

The exponentially growing COVID-19 time series data can be modelled and studied using Auto Regressive Integrated Moving Average (ARIMA) model[1] This model can be used to do short term prediction[2]. The data is taken from the Johns Hopkins university (https://gisanddata.maps.arcgis.com/apps/opsdashboard/index.html) are useful because they can provide a forecast for COVID-2019 pandemic to effectively control the spread of this highly infectious disease in India. Depending on the predictions, the government officials should adapt aggressive interventions to control this exponential growth [3] of this rapid infectious disease and curtail the COVID-19 pandemic. The updation of Johns Hopkins university on a daily basis is considered and the data for India till 14 ^th^ April 2020 is considered for this analysis and a time-series database was created in Excel. Exploratory data analysis of the data was performed to predict on a short term prediction of confirmed cases of COVID-19 in India for the next 10 days is predicted effectively.

## 2. Methodology

ARIMA forecasts on its previous past values and there are three distinct integers (p, d, q) that are used to parametrize ARIMA models. The three parameters account for seasonality, trend, and noise in datasets are denoted with the notation ARIMA(p, d, q). In the model, ***p*** is the auto-regressive part of the model and incorporates the effect of past values in the model. ***d*** is the integrated part of the mode and incorporates the amount of differencing to apply to the time series. The parameter ***q*** is the moving average parameter that allows to set the error of the proposed model as a linear combination of the error values observed at previous time points in the past. Our goal is to that optimizes the metric of interest. The experiment is carried out in R Programming. The Fig 5. Plots the cumulative cases from the John Hopkins University dataset for India.

The augmented Dickey-Fuller (ADF) test is a formal statistical test done to ensure stationarity. In ARIMA modeling using R the univariate data is converted into time series data format. The graph follows an overall upward trend with some outliers in terms of sudden lower values. The Augmented Dickey Fuller Test (ADF) is unit root test for stationarity. Since the data is not stationary, the data is differenced and is computed by the differences between consecutive observations.

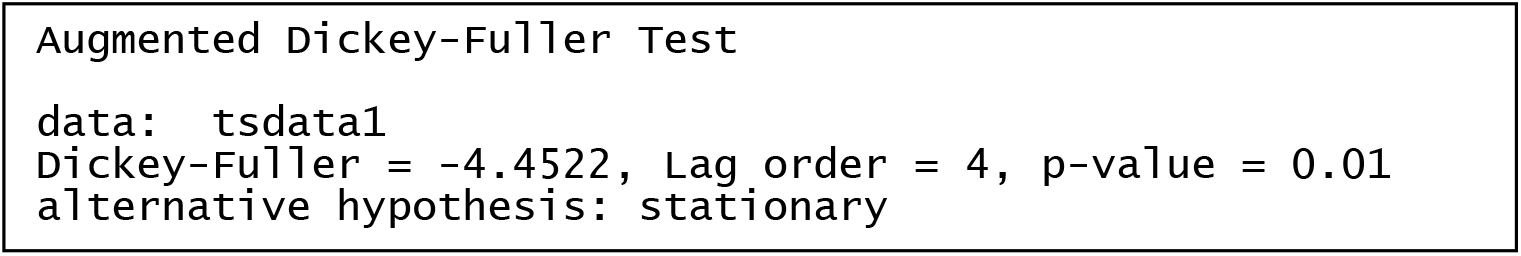

Since the p-value after differencing is 0.01 and is less than 0.05 the null hypothesis is rejected and the data does not have a unit root and is stationary. The time series after the data is removed of its non stationarity is given in the Fig 1.

**Fig 1.**
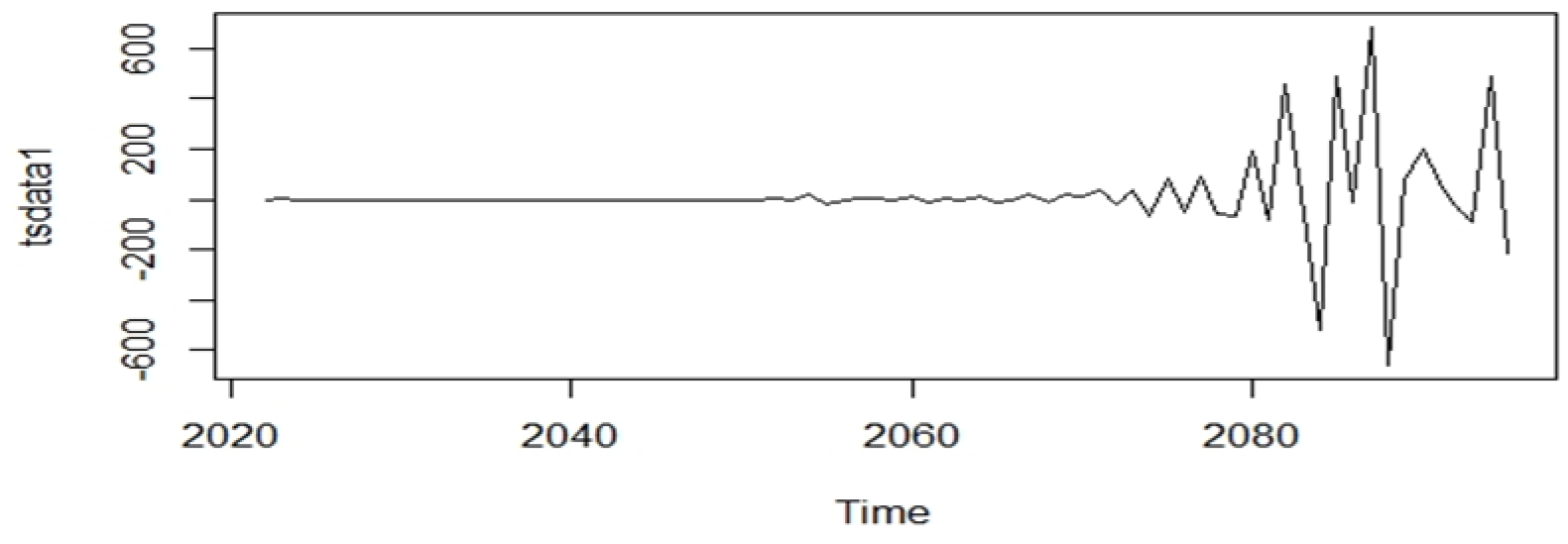
Non Stationarity removed Time series.

The autocorrelation function gives the autocorrelation at all possible lags. The autocorrelation at lag 0 is included by default which always takes the value 1 that represents the correlation between the data and themselves. The ACF and the PACF is given in Fig.2

**Fig 2.**
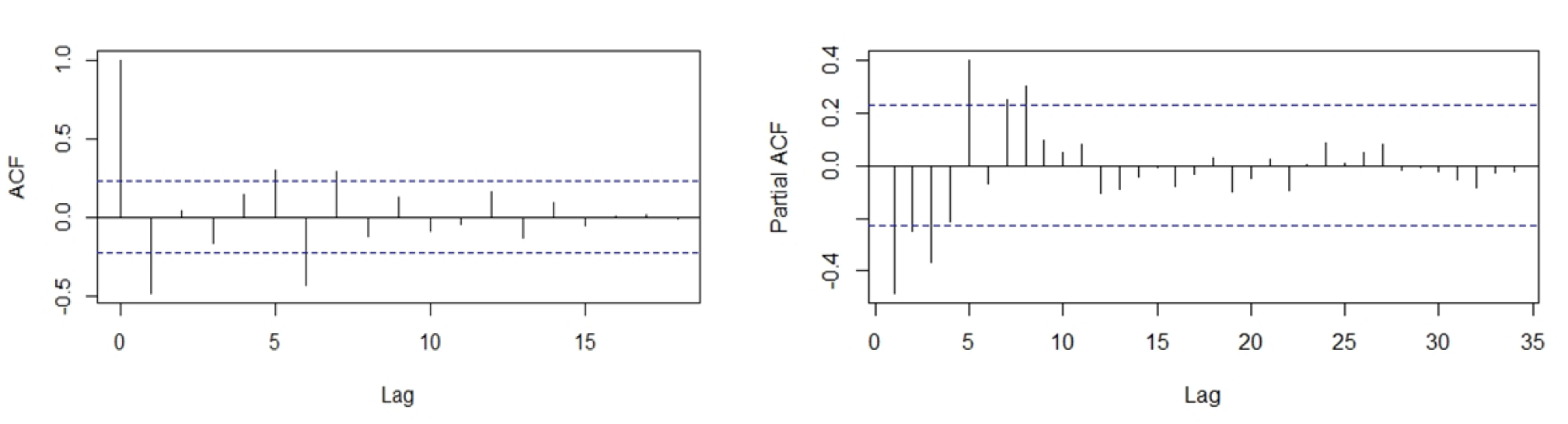
ACF and PACF Factors.

**Fig 3.**
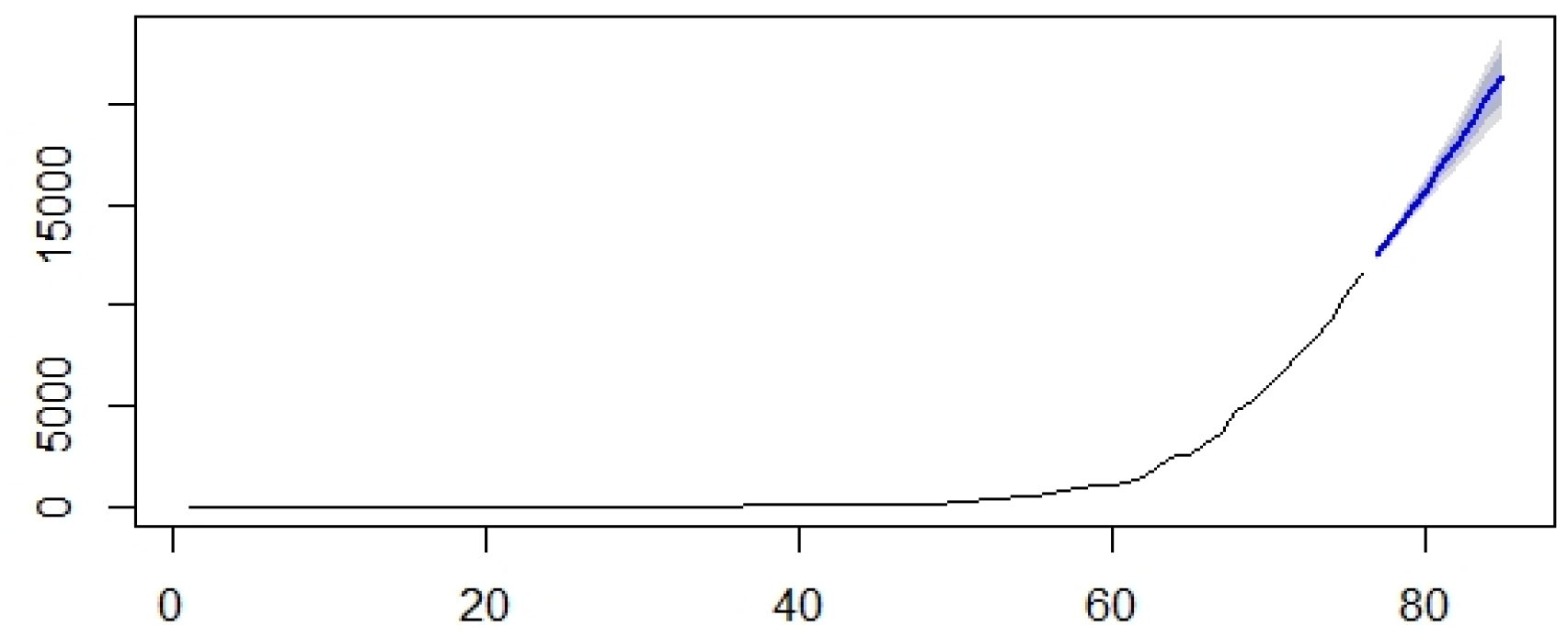
ARIMA Forecast of confirmed cases of COVID-19 in India.

The best fit model is selected based on Akaike Information Criterion (AIC) value of the model. The idea is to choose a model with minimum AIC and BIC values.The best model is ARIMA(1,2,2) with the AIC value of 932 and the the BIC value of 941is fitted using the auto.arima() function.

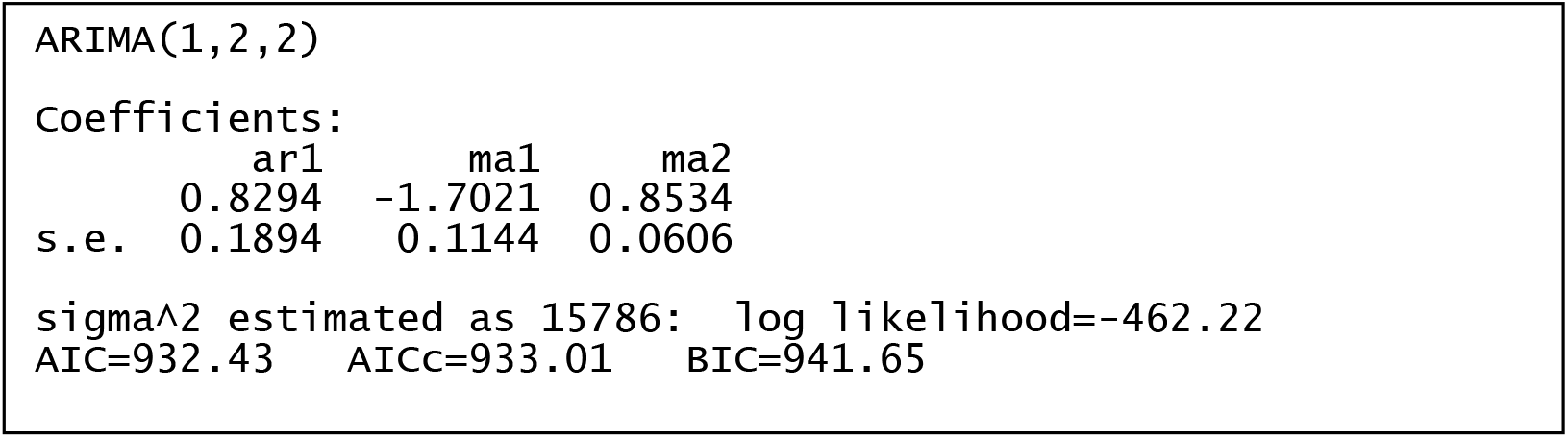

The forecast for the next 10 days is given below,

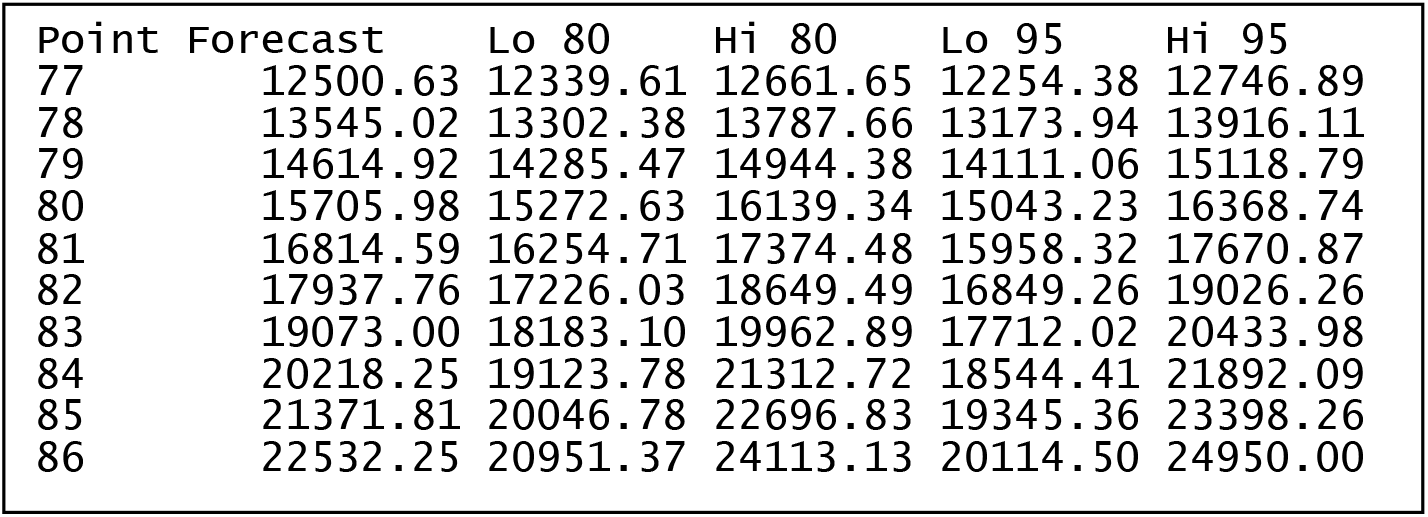

The forecast of the Confirmed Cases for the next 10 days, that is, until 24^th^ April 2020. The Forecast reaches 22532 on the 86^th^ day, that is, by 24^th^ April 2020 if proper social distancing and other measures are not followed. The Figure plot shows the predicted cases. The blue line represents the forecast and the silver shade around it represents the confidence interval.

This Ljung-Box Q test assess the overall randomness based on a number of lags, and is therefore a portmanteau test. It is applied to the residuals of a fitted ARIMA model, not the original series, and in such applications the hypothesis actually being tested is that the residuals from the ARIMA model have no autocorrelation.

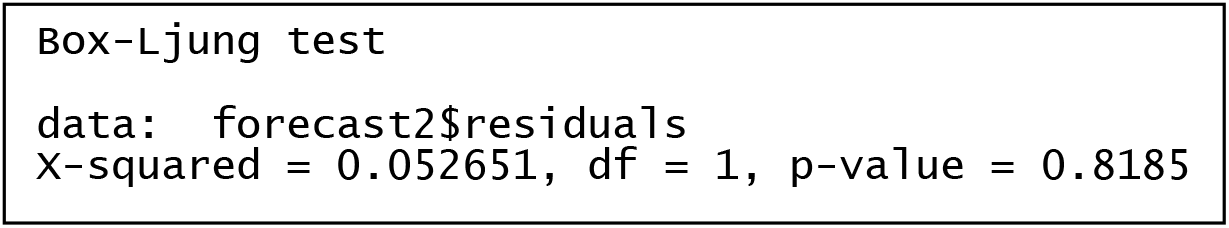

The different metrics to evaluate the model is given in the following Table 1. That shows significant performance values.

**Table 1.**
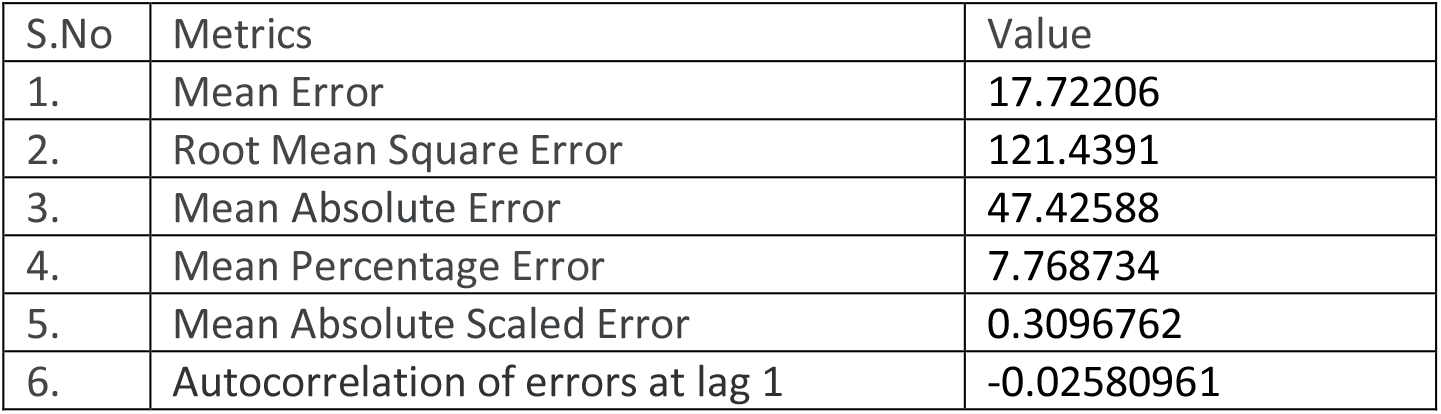
Performance Metrics of ARIMA Model

| S.No | Metrics | Value |
| --- | --- | --- |
| 1. | Mean Error | 17.72206 |
| 2. | Root Mean Square Error | 121.4391 |
| 3. | Mean Absolute Error | 47.42588 |
| 4. | Mean Percentage Error | 7.768734 |
| 5. | Mean Absolute Scaled Error | 0.3096762 |
| 6. | Autocorrelation of errors at lag 1 | -0.02580961 |

This model assume that the population of COVID-19 and the infected but not yet isolated population have the same contact rate. The factors like lock down, social distancing, wearing of masks and usage of sanitizers are not considered while modelling and predicting the confirmed cases of COVID-19.Also the continuous release of the epidemic data there might be changes in the spread of of COVID-19 among the population.

## Data Availability

https://gisanddata.maps.arcgis.com/apps/opsdashboard/index.html

